# Impact of SARS-Cov-2 on Clinical Trial Unit Staff: The EPIC Observational Study

**DOI:** 10.1101/2022.07.18.22277769

**Authors:** Peter Phiri, Lucy Yardley, Kathryn Elliot, Katharine Barnard-Kelly, Vanessa Raymont, Shanaya Rathod, Jintong Hu, Heitor Cavalini, Jian Qing Shi, Gayathri Delanerolle

## Abstract

**Introduction:** Clinical Trials Units (CTUs) are a key component of delivering non-commercial and commercial clinical research globally. Within the UK, CTUs are seen as a specialist and independent entity available to all researchers requiring support to setup, conduct and deliver clinical trials. Therefore, an involvement of a CTU is highly recommended by national regulators and positively accepted by funders, especially for drug and/or medical device and/or complex intervention trials.

**Aim:** This study aims to determine the challenges associated with the management of Covid-19 research managed via the CTU workforce, including the challenges associated with quality assurance, trial setup and data management. Additionally, this study will explore the by-stander effect on trial staff by way of evaluating the mental and physical health impact.

**Methods/ Design:** This is a mixed methods study. An online novel questionnaire survey study will be conducted among the UK CTU workforce. Quantitative data will be collected using the Qualtrics XM platform. We aim to recruit up to 1,500 CTU staff across the UK workforce. A subgroup sample will be randomly invited to take part in semi-structured interviews. Therefore, this survey will generate both quantitative and qualitative data inclusive of demographic data.

**Results:** The findings will inform current initiatives and identify key themes for prioritising in further research to develop robust approaches to support CTU staff, including the development of a start-re-start framework for CTUs for any future pandemics relevant to developing and delivering communicable diseases and non-communicable diseases-based research.

**Strengths/Limitations:** The validation of the EPIC impact questionnaire used qualitative and quantitative methods which is a strength of the study. However, the study has a single timepoint to obtain data with the secondary outcome measures to be completed at two timepoints as this is an exploratory study attempting to obtain a wider data pool.

## Introduction

Covid-19 has presented challenges for staff working within CTUs. Within most organisations, some CTU staff are shared between clinical and scientific roles. Therefore, most CTU staff have had to adapt rapidly to the use of a variety of non-standard procedures. As a result of this, CTU staff have worked under significant pressure. Therefore, the trial community’s mental wellbeing is key to ensuring they are able to continue to share their experiences and expertise in the future with innovative practices to support the continuous evidence generate to improve clinical practices in the future. To achieve this, it would be useful to undertake a survey to help obtain the views of the trial community within CTUs in the UK, to ensure, the relevant support mechanisms could be put in place for the future.

Previous studies on the outbreaks of infectious diseases such as severe acute respiratory syndrome (SARS) and Middle East respiratory syndrome (MERS), have consistently showed adverse psychological impacts on health care workers although this was not evaluated in the context of CTUs. These impacts include high levels of anxiety, depression, and stress that resulted in many healthcare workers meeting the diagnosis of a post-traumatic stress disorder [1-2]. These findings are supported by a recent study exploring the impact of covid-19 [3]. This showed a considerable proportion of health care workers in China experienced a high level of anxiety and depression, insomnia and psychological distress. Healthcare workers in locations affected by pandemics are prone to symptoms of anxiety and depression [4]. While informative, it is unclear whether these findings are generalizable to UK CTU workforce.

The CTU workforce in the UK is approximated around 25,000 although some staff have shared roles with multiple departments spanning across academic and NHS organisations. Therefore, it is vital to obtain all staff associated with CTUs to discuss about their experiences during this pandemic.

### Challenges associated with the management of Covid-19 trials

Bhatt [5] indicated important issues around managing covid-19 clinical trials associated with scientific and ethical principles. The clinical and scientific issues are primarily associated with the investigational medicinal products (IMPs) or interventions as well as study design, assessments of efficacy and safety implications, securing informed consent, sample size and publication. Similarly Delanerolle et al [6] indicate the ethical implications associated with the staff delivering this work that is not reviewed when research ethics are assessed when global health hazards such as a pandemic takes priority. Davies et al [7] also pointed out the lockdowns, site closures and recruitment from hospital sites themselves have caused interruptions in addition to challenges in receiving supply chains of IMPs. Staff associated with each of these steps had the risk of being infected with Covid-19. The Indian Society for Clinical Research (ISCR) for example published a position paper providing guidance to both frontline clinical research professionals and sponsors to measure ongoing activities as well as a method to resume site activities. Additionally, the NIHR also focused in providing guidance to research organisations within the UK although this was put together in haste without any involvement with clinicians and patients. Evidence based approaches are vital to use when developing such frameworks in the future.

### Challenges associated with Covid-19 in terms of quality assurance

The quality issues associated with covid-19 trials and its implications requires further research to assess the extent of any quality assurance issues. However, based on publications and viewpoints published thus far, it appears there is a lack of high-quality methods being opted for in order to meet the urgent demands of the population. Shiely and colleagues [8] conducted a study to understand the management of clinical trials to offer practical advice, although quality assurance was not one of their focuses. Regulatory authorities such as the European Medicines Agency (EMA), the medicines and healthcare products regulatory agency (MHRA), the food and drug administration (FDA) have all provided various guidance tools to direct clinical trial staff although, navigating around these have been cumbersome for many due to a variety of reasons including differences in healthcare systems and infrastructure.

### Healthcare professionals and Burnout

In the UK, the provision of NHS care is rendered by its workforce, comprising over 1,236,000 professionally qualified clinical staff in England [9]. As such, the quality of the NHS care delivery is substantially dependent on the availability and performance of its health and social care workforce [28-29]. Approximately, 1 in 5 (20.7%) of this workforce were from a BAME background [10]. The Management of Health and Safety at Work Regulations (1999 as amended) requires employers to ensure the work environment is, as far as reasonably practicable, safe and without risks to health.

Burnout amongst healthcare professionals is a key challenge affecting healthcare practice, safety and quality of care. Khasne and colleagues [11], Alsulimani et al [12], Mehta and colleagues [13] and Tan et al [14] reported burnout amongst healthcare professionals which sets precedence to explore CTU staff who are often a combination of dedicated and shared staff. This also means that there is currently a gap in the literature to identify the challenges associated with the CTU workforce.

### Value to NHS

The clinical trial workforce provides a vital component for the NHS and public welfare. CTUs are pockets of multidimensional skills and the workforce is consistent of clinical and scientific experiences as well as expertise. Firstly, evidence shows that Trusts which are more intensively involved in research have better clinical outcomes, even for patients not directly enrolled in clinical trials. Participant experience surveys run regularly by the NIHR CRN show that 90% of patients have a good or better experience of research. The majority take part through altruism. They want future patients to have improved treatments and for knowledge of their condition to advance. They also often receive access to novel treatments or better monitoring of their condition. Research-active organisations encourage their clinicians to be aware of current thinking and best practice for all the care they provide.

## Methods and Analysis

### Methods

To achieve these objectives, a cross-sectional observational study has been designed to be delivered digitally. Quantitative data will be collected through an online survey questionnaire using the Qualtrics Core XM platform to understand challenges the trial community within CTUs faced in addition to the mental health and wellbeing aspects endeavoured by staff.

#### Hypothesis

CTU staff have provided extensive support to deliver clinical research /trials during pandemics that may have had an impact on their wellbeing the same healthcare professionals. Infrastructural and Operational requirements would vary across CTUs as most of these are within higher education institutions (HEIs) and/or the NHS. Therefore, improvements and perhaps unified approaches to better prepare to deliver future clinical trials/research in the event of a future pandemic should be considered. To do this, sufficient operational and infrastructural information from all the staffing groups in CTUs would be useful.

##### Primary objectives

1. Determine the challenges associated with trial management for covid-19 studies
2. Determine the challenges associated with quality assurance for covid-19 studies

##### Secondary objectives

1. Determine the challenges associated with for non-covid related studies in terms of trial management
2. Determine the challenges associated with covid-studies in terms of quality assurance
3. Determine the wellbeing challenges whilst working during the lockdown periods

##### Exploratory objectives

1. Determine any local framework(s) used to deliver covid-19 studies remotely
2. Determine any local framework(s) used to deliver non-covid studies remotely
3. Determine any *continuous improvement plan* used to demonstrate a formalised best practice guide whilst working remotely and delivery covid-19 studies

Recommendations based on these findings will be made to inform improved future preparedness in supporting CTU workforce delivering to pandemic and non-pandemic research.

#### Study Participants

All staff working in CTUs in the UK would be able to take part in this study through the online survey. Our sample size would be 1,500 although this may exceed depending on the uptake to the survey. The survey would be deployed online via the NIHR, UKTMN, social media, MRC and the UK CRC. All participants will be required to complete the survey at a single time point for the primary and exploratory outcome measures, with the secondary outcome measures completed at two time points. An online Qualtrics Core XM platform survey will approximately take 10 -15 minutes to complete following the completion of consent.

Participants will be invited to participate in the study via multiple media sources including intranet, email invites and social media, newsletters and communication campaigns supported by their organisations. The survey will be deployed online via the NIHR, UKTMN, Social Media, MRC and the UK CRC. Informed consent will be given on the first page of the survey. If participants agree to take part in the study by clicking the consent boxes, they will be able to proceed to the online survey questions. However, if they do not agree to take part they will not be able to enter the survey.

##### Inclusion Criteria

- ≥18 years.
- Any gender
- CTU staff working the UK
- Have access to a smartphone, tablet or computer to be able to complete the survey online.
- Willing to give informed e-consent

##### Exclusion Criteria

- <18 years
- Those unwilling to participate in the study

### Quantitative

#### EPIC Pandemic questionnaire design

The questionnaire comprises of 99 questions with 4 dimensions and designed to be completed online using the online Qualtrics XM platform.

##### PART A

Participant demographic characteristics, this section will ascertain participant personal information such as age, gender, ethnicity, marital status, job title, healthcare professional status, employment sector, setting, nationality and religion, level of education. For the BAME groups the Vancouver Index of Acculturation (VIA) will be used to measure levels of acculturation to the British culture.

##### PART B

The core questions about and mental health and wellbeing will be measured using the validated HADS, the Pandemic Index Scale, and the Insomnia Severity Index Scale.

##### PART C

Operational including flexibility to working remotely, involvement in covid related research at set up to delivery; use of covid-19 associated risk assessment procedures; productivity and confidence in career progression; Perceptions of increased risk, adverse conditions including working hours. Validated measures will include the General Self-Efficacy Scale and the Burnout Assessment Tool (BAT-12) measure. PPE availability, training and usage

##### PART D

Experiences of everyday discrimination will be measured using the Everyday Discrimination Scale (EDS) and Agency for Healthcare Research and Quality (AHRQ).

Participants will be allowed to omit questions they do not want to complete. The survey is anonymous. The survey will take approximately 15 minutes to complete.

#### Primary and secondary outcome measures

Primary outcome: Determinants of challenges associated with trial management, quality assurance of covid-19 trials and non-covid trials, including wellbeing.

#### General Self Efficacy Scale (GSE)

The GSE was originally developed in Germany and has been adapted to 28 languages [15]. Numerous studies have demonstrated the GSE to have High reliability, stability, and construct validity [16-17]. The scale includes only one global dimension measured through 10 items. Participants respond to items such as “Thanks to my resourcefulness, I can handle unforeseen situations” using a 4-point Likert scale ranging from ‘not at all true’ through to ‘exactly true’.

#### Hospital Anxiety and Depression Scale (HADS)

The HADS is a 14-item self-report measure of anxiety and depression. It is scored on a 4-point Likert scale [18, 21]. It has good internal consistency and good concurrent validity [19-20]. This measure was designed for a non-clinical population but has recently been validated for a mental health population and the reliability estimates have shown that the Cronbach’s alpha for HADS (all items), HADS-Anxiety subscale (HADS-A) and HADS-Depression subscale (HADS-D) was .91, .90 and .80, respectively [21]. The maximum score is 21 for depression and 21 for anxiety. A total score of 11 or higher indicates the probable presence of the mood disorders with a score of 8 to 10 being just suggestive of the presence of the respective state.

#### Burnout Assessment Tool (BAT-12)

The BAT-12 a short version self-reported questionnaire consisting of 12 items in four domains namely, exhaustion, mental distance, cognitive impairment and emotional impairment [22]. Each statement is scored on a range of 1 (never) to 5 (Always).

#### Insomnia Severity Index (ISI)

The Insomnia Severity Index (ISI) is a 7 item self-reported measure of sleep quality assessing the nature, severity and impact of insomnia within the last month [23]. Individual items are scored on a 5-point Likert scale ranging from 0(no problem) to 4 (being very severe problem) with a total maximum score of 28. A score of (0–7) is indicative of no insomnia, (8–14) indicative of a sub-threshold insomnia, (14–20) moderate insomnia and (22,28,29) is indicative of severe insomnia.

#### Pandemic Stress Index (PSI)

**Table 1.**
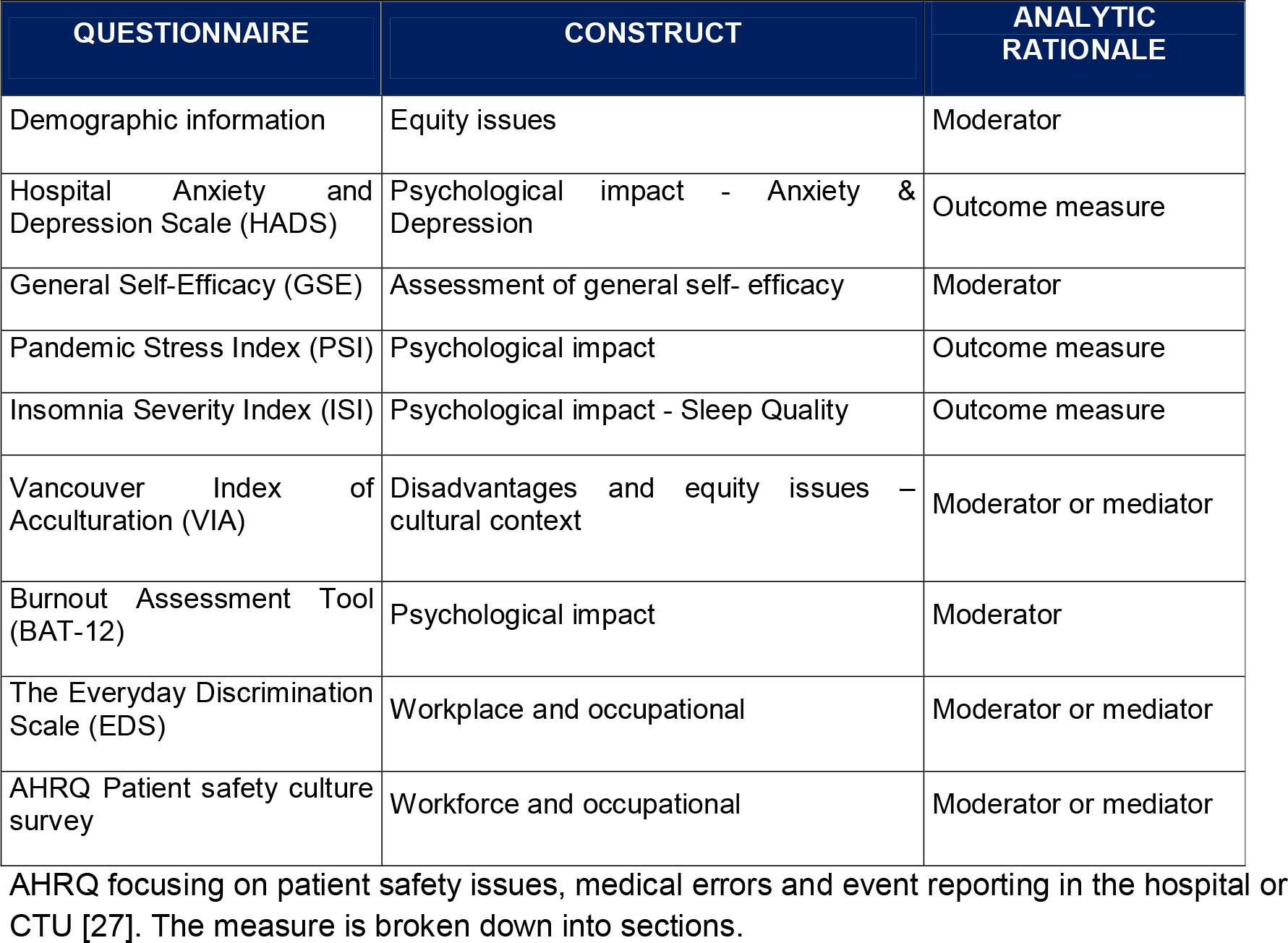
Table of Questionnaires

*AHRQ Patient safety culture survey*
| QUESTIONNAIRE | CONSTRUCT | ANALYTIC RATIONALE |
| --- | --- | --- |
| Demographic information | Equity issues | Moderator |
| Hospital Anxiety and Depression Scale (HADS) | Psychological impact - Anxiety & Depression | Outcome measure |
| General Self-Efficacy (GSE) | Assessment of general self- efficacy | Moderator |
| Pandemic Stress Index (PSI) | Psychological impact | Outcome measure |
| Insomnia Severity Index (ISI) | Psychological impact - Sleep Quality | Outcome measure |
| Vancouver Index of Acculturation (VIA) | Disadvantages and equity issues – cultural context | Moderator or mediator |
| Burnout Assessment Tool (BAT-12) | Psychological impact | Moderator |
| The Everyday Discrimination Scale (EDS) | Workplace and occupational | Moderator or mediator |
| AHRQ Patient safety culture survey | Workforce and occupational | Moderator or mediator |

The PSI is a descriptive measure that assesses self-reported behavioural, psychosocial, and medical experiences during COVID-19 comprising three items [24]. The first item asks, “What are you doing/did you do during COVID-19?” and instructs participant to check all items that apply. Item 2 asks participants to rate the impact of COVID-19 on their day-to-day life on Likert scale 0 (being not at all) and 5 (being extremely). Item three, asks participants, “Which of the following are you experiencing during COVID-19?” and provides a list of potential experiences to choose all that apply, e.g. “more depression, more sleep, and more anxiety.”

#### The Vancouver Index of Acculturation

VIA is a 20-item scale measuring level of acculturation to the host culture, British culture. Likert scale ranges from 1, disagree to 9 agree [25].

#### The Everyday Discrimination Scale

The EDS it is used as a measure of subjective experiences of daily discrimination against the minority population [26]. This measure contains nine elements that assess the person’s daily life, followed by a follow-up question about what the person believes was the reason for that daily discrimination.

#### AHRQ Patient safety culture survey

##### Early Discontinuation/Withdrawal of Participants

If a participant who has given informed consent decides during the course of the study to discontinue or withdraw their participation prior to submitting their questionnaire they will be withdrawn from the study. However once submitted we will not be able to identify them unless they have given their contact details then they will be withdrawn. Identifiable data already collected with consent would be retained and used in the study. No further data would be collected or any other research procedures carried out in relation to the participant.

##### Data management

The online survey data will be collected using the Qualtrics Core XM platform. Survey data will be exported from the Qualtrics Core XM platform to the statistical software packages SPSS and STATA for graphical presentation and analysis.

##### Potential Risks

The risks associated in taking part are very minimal. Taking part may make participants think more. The team does not anticipate any potential adverse effects, pain, discomfort, distress, intrusion, inconvenience or changes to lifestyle of participants through the survey. However, should any participant experience distress they will be able to access workplace options support services, occupational health services and/or supervision from employer organisation. Furthermore the survey will signpost staff to contact their general practitioner or NHS 111 service. The survey will take approximately 15 minutes; therefore this will be of minimum burden to participants. Access to online survey means that the participants will be able to complete the questionnaires in their own time without the added responsibility of returning their responses to the researcher.

##### Data Security

The Southern Health NHS Foundation Trust Qualtrics account is password protected by a high-end firewall system. Personal data, that is, email addresses or telephone numbers will only be collected for participants who wish to participate in the qualitative subgroup interviews. All data will be collected in a secure password protected online Qualtrics Core XM platform. Access to systems is severely restricted to specific individuals, whose access is monitored and audited for compliance.

Data exported from Qualtrics Core XM platform will be anonymous, stored and managed in password protected files in a password protected computer. Only members of the research team will know the passwords and will therefore be able to access the electronic data. Study documentation will be archived in accordance with guidelines for Good Clinical Practice and in NHS approved, secure and adequate archiving facility. Research personnel will keep information relevant to the study for up to 10 years, and then will be destroyed.

All qualitative interviews will be conducted via a secure online facility (password protected platform such as zoom teleconference), audio-recorded, transcribed by researchers in the study team. Transcriptions will be deidentified and uploaded to NVivo software version 11.2 for data coding/retrieval.

##### Definition of End of Study

The end of the study is defined as the completion of the survey and qualitative interview data collected from the last participant.

### Qualitative

All interviews will be conducted via secure online facility (password protected zoom teleconference), audio-recorded, transcribed in full, and early interviews will be reviewed by the research team to determine whether any alterations to the topic guides need to be made.

Data collection and analysis will be integrated. A process based on Framework methodology will be used to analyze the data:

- Development of a coding frame based on identified key themes
- Detailed indexing (coding) of transcripts
- Charting to organize and summarize data
- A detailed review of the charted data to facilitate interpretation

A semi-structured interview script will be developed to guide the interviews. (See appendix 1 for the interview guide. Interviews with 5-10 CTU staff will be conducted to ensure capture of all relevant key determinants. If data saturation is reached, interview capacity will be shifted to other groups to facilitate greater exploration where appropriate. It is anticipated that interviews will take approximately 45 minutes, however this will be guided by each participant.

### Analysis

#### Quantitative analysis

- Descriptive statistics will be presented as either means (SD) or median (IQR) for continuous variables according to the distribution of data
- Descriptive statistics for categorical data will be presented as frequencies and proportions.
- Parametric or non-parametric statistics will be chosen depending on the distribution of the data. And if appropriate data transformation will be done for carrying out parametric analyses.
- Data will be analysed using one-way analysis of variance (ANOVA) and t-tests to assess associations between demographic characteristics
- Where data is not normally distributed, non-parametric techniques (Kruskal-Wallis test and Mann-Whitney U-test) will be utilised.
- Chi-square or Fisher’s exact test will be used for categorical variables and any associations between demographic data and responses related to mental health wellbeing questions.
- Statistical significance is assumed at p<0.05.
- Pearson’s correlation analysis to measure strength of linear relationship between variables. Also if appropriate non-parametric correlation will be explored if data are skewed.
- Multiple regression analysis will be carried out for continuous dependent variables with adjustment for appropriate confounders / covariates. As the survey is multi-national we shall employ multi-level models also.
- Logistic regression analysis will be carried out for categorical dependent variables with adjustment for appropriate confounders / covariates.
- The statistical software packages SPSS and STATA will be used for graphical presentation and data analysis. Additionally statistical package R will be considered for graphical output if appropriate.

#### Qualitative

The initial coding frame will be agreed after review of a selection of transcripts by the study qualitative researchers however, a constant comparative approach will be adopted whereby the coding frame will be open to revision during the detailed coding and charting stages of the process of analysis.

Interviews will be transcribed by the researchers. Transcripts will then be reviewed for accuracy, de-identified, and uploaded to NVivo software version 11.2 for data coding/retrieval. Transcripts will be analyzed using thematic and content analysis. Two experienced qualitative researchers will independently review transcripts and conduct analyses. A coding framework will be developed to capture key themes with each coded theme subjected to detailed analyses to identify subthemes and illustrative quotes. After the initial two investigators establish the codebook, the codes will be applied by the members of the research team. It is anticipated that this research will:

- Play a vital role in aiding interpretation of the quantitative data
- Develop insights of CTU staff’s experiences of COVID-19 pandemic to determine the challenges associated with trial management, quality assurance and impact on wellbeing.
- Provide recommendations

#### Potential Bias

This is a self-reported questionnaire; one likely bias may be social desirability – but this would be minimum as the survey is anonymous. We anticipate low bias in demographic information part of the survey. The established questionnaires that we have chosen have already been investigated for validity and reliability in many settings. Thus overall we expect low level of bias.

#### Sample Size Determination

There are 53 UK CRC registered CTUs although an excess of 150 exist in various shapes and forms scattered around academic and NHS organisations. We envisage at least 400 participants would suffice as the sample size.

## Data Availability

All data produced in the present study are available upon reasonable request to the authors

## Acknowledgements

The authors acknowledge support from Southern Health NHS Foundation Trust.

## Ethics and Dissemination

HRA REC approval (21/HRA/2348) was obtained prior to the study initiation. All resulting publications would be disseminated in a peer reviewed open access journal. Additionally, the findings will be disseminated via the UKCRC and social media channels.

## Author contributions

GD and PP developed the study protocol and embedded this within the EPIC project’s work-package 3. The Chief Investigator of this study is PP and Principal Investigator GD. GD, JT and JQS designed and completed the statistical analysis. The qualitative analysis was completed by KBK, PP and GD. All authors critically appraised and commented on previous versions of the manuscript. All authors read and approved the final manuscript.

## Availability of data and material

The authors will consider sharing the dataset gathered upon receipt of reasonable requests.

## Code availability

The authors will consider sharing the dataset gathered upon receipt of reasonable requests.

